# Outcomes and Risk Factors for Postoperative Vasoplegia Following Implant of Continuous-Flow Left Ventricular Assist Devices

**DOI:** 10.1101/2024.05.13.24307313

**Authors:** Jonathan C. Wright, Jameson G. Wilbur, Sujal Modi, Mahmoud Abdel-Rasoul, Bryan A. Whitson, Asvin M. Ganapathi, Sakima Smith

## Abstract

Refractory postoperative vasoplegia following cardiac surgery and left ventricular assist device (LVAD) implantation predicts poor outcomes. We aimed to investigate outcomes and predictors of postoperative vasoplegia. We retrospectively reviewed a single-center cohort of 190 patients who received LVADs from January 2015 through March 2022 at The Ohio State Wexner Medical Center. Primary outcomes included duration of ICU stay, development of right heart failure (RHF), and mortality. Secondary outcomes included pre-implant medications and post-implant blood products. Vasoplegia was defined as physician documentation of vasoplegia with patients requiring ≥1 intravenous vasopressors within 48-hours following LVAD implantation to maintain a mean arterial pressure >65 mmHg for >24 hours. Overall, 55 (29%) patients developed vasoplegia following LVAD implantation. Our sample was stratified into two cohorts: patients with vasoplegia and without vasoplegia. Baseline characteristics, LVAD indication, creatinine clearance, and comorbidities did not vary significantly between cohorts. Patients without vasoplegia were more likely to have prior implantable cardiac defibrillators (p = 0.02) and were more commonly prescribed pre-operative inotropes (p = 0.03). Blood-product administration within the first 48-hours postoperatively did not differ. While patients with vasoplegia required a longer ICU stay post-op (5 vs 7 days, p = 0.03), there were no significant differences in development of RHF (39% vs 44%), 30-day (7% vs 13%), 1-year (31% vs 29%), and 2-year mortality (39% vs 33%) in patients without and with vasoplegia, respectively. Overall, our vasoplegia rates were consistent with previous literature (29%) and vasoplegia did not confer inferior outcomes for mortality or developing RHF.

## Introduction

Vasoplegia syndrome or vasodilatory shock is a clinical condition characterized by decreased systemic vascular resistance and severe systemic hypotension. This manifests as shock refractory to fluid resuscitation and vasopressors, despite a normal or even increased cardiac output^1,2^. Vasoplegia is a common postoperative complication of cardiac surgeries with reported incidence varying widely, but occurring in as high as half of all patients^3–6^.

Treatment guidelines for vasoplegia are lacking. Much of the current treatment is extrapolated from septic shock which presents with similar refractory vasodilation. Typically, vasoplegic patients are treated with a combination of therapies, with the cornerstone involving vasoactive agents such as norepinephrine. Newer therapies are targeted at the various biological pathways thought to be involved in the pathophysiology of vasoplegia such as the arginine-vasopressin system (vasopressin), renin-angiotensin-aldosterone system (angiotensin II), and modulators of nitric oxide and other inflammatory mediators (methylene blue, hydroxocobalamin, vitamin C, and corticosteroids)^3^. However, studies investigating methylene blue failed to show improvement in clinical outcomes^7^.

Specifically in left ventricular assist devices (LVAD), vasoplegia is a major complication following implantation^8^. Postoperative vasoplegia has been associated with poor outcomes in LVAD patients, including major bleeding, increased length of hospital stay, respiratory failure, and right heart failure^6,8^. Furthermore, studies which stratified patients by vasoplegia severity found a higher risk of early mortality in those considered to have more “severe” vasoplegia^8^. Despite this, some smaller studies suggest that LVAD patients who develop vasoplegia post-operatively have equivalent long-term outcomes to those who do not develop vasoplegia^9^.

Despite its clinical burden, few studies exist exploring risk factors for vasoplegia in LVAD recipients. One study found elevated creatinine clearance and pre-operative beta-blocker use were associated with decreased risk of vasoplegia in patients undergoing heart failure related surgeries, including LVAD implantation^6^. Other risk factors identified have included preoperative Interagency Registry of Mechanically Assisted Circulatory Support (INTERMACS) profile, central venous pressure, systolic blood pressure, and intraoperative cardiopulmonary bypass time^8^.

In this present study, one of the largest of its kind, the purpose was twofold. First, we aimed to compare outcomes in those who developed vasoplegia perioperatively versus those who did not. Secondly, we aimed to investigate risk factors associated with the development of vasoplegia in the perioperative period following LVAD implantation.

## Methods

This was a retrospective cohort study. Prior to data collection the study protocol was evaluated and approved by the institutional review board at The Ohio State University Medical Center (#2019H0328).

### Setting and Participants

We included all patients who had undergone LVAD implantation at The Ohio State University Wexner Medical Center between January of 2015 and March of 2022. LVAD devices included in our study were HeartMate II (n = 69, 36.3%), HeartMate III (n = 45, 23.7%), and HeartWare Ventricular Assist Device (HVAD) (n = 76, 40.0%). The exclusion criteria were patients who had their LVAD implanted at an outside hospital and transferred to The Ohio State University Wexner Medical Center for higher level of care.

### Variables

Early right heart failure (RHF) was defined by 2014 INTERMACS definition of right heart failure within 30 days of LVAD implantation and required both documentation of elevated central venous pressure (CVP) and clinical manifestations of elevated CVP^10^. Documentation of elevated CVP included: direct measurement of right atrial pressure > 16 mmHg, significantly dilated inferior vena cava with absence of inspiratory variation on echocardiography, or clinical findings of elevated jugular venous distension at least halfway up the neck in an upright patient^10^. Clinical manifestations of elevated central venous pressure included peripheral edema (>2+ either new or unresolved), ascites, palpable hepatomegaly on physical examination or imaging, laboratory evidence of worsening hepatic dysfunction (total bilirubin > 2.0 mg/dl), or worsening renal dysfunction (creatinine > 2.0 mg/dl)^10^.

Late RHF was defined as a patient who had both clinical symptoms of right ventricular function and required hospitalization for inotropic support greater than 30 days after implant but within 1 year of implant^11^. Clinical symptoms of right ventricular dysfunction included hepatic congestion, peripheral edema, and jugular venous distension^11^. The readmission must have occurred more than 30 days after discharge from the index LVAD implantation in an effort to avoid capturing cases defined as early RHF^11^. Patients with early RHF were excluded from those with late RHF.

Currently, there is no validated definition for vasoplegia and definitions from prior studies varied widely. Such studies have turned towards objective measurements such as vasodilation criterion based on mean arterial pressure, hemodynamic criterion based on cardiac index, preload criterion based on central venous pressure, and vasopressor use^8,12–25^. The most common factors included in definitions of vasoplegia include use of at least one vasopressor to maintain mean arterial pressures >50-70 in the first 24-48 hours post-device implantation, in the absence of other explainable causes for persistent hypotension (cardiogenic shock, sepsis, etc.). As such, to capture as many patients as possible who were treated for suspected vasoplegia, our study defined vasoplegia as provider documentation in the electronic health record as vasoplegia as the most likely cause of persistent hypotension (MAP <65 mmHg) requiring the use of at least one vasopressor in the first 48-hours after device implantation in the absence of other causes for persistent hypotension.

For all patients our primary endpoints were duration of ICU stay, duration of hospitalization, development of early or late RHF, and 30 day, 1-year, and 2-year mortality. Secondary endpoints included post-operative blood product administration and pre-implant medications.

### Data Sources

All patient data including demographic information, procedural data, and outcomes data were collected retrospectively from our hospital’s electronic health record.

### Statistical Methods

We used both parametric and non-parametric statistics to summarize and compare our cohorts. Discrete ordinal variables, such as length of ICU stay in days, were compared using a Kruskal-Wallis analysis of variance. Nominal variables, such as proportion of patients diagnosed with hypertension, were compared using a Chi-squared test. Continuous variables, such as age, BMI and creatinine, were analyzed using one-way analysis of variance. Statistical significance threshold p < 0.05. All statistical analyses were performed by an independent statistician.

## Results

### Participants

The initial query with ICD-9 code V43.21 and ICD-10 code Z95.811 which identified 195 patients who received LVAD implantation during the study period. Three patients were excluded due to LVAD implantation at an outside hospital and two patients were excluded due to Total Artificial Heart implantation for a final study size of 190 patients. Participant follow-up time ranged from 1 day to 8.0 years with a median follow-up time of 2.47 years with an interquartile range of 0.55 – 5.05 years.

### Characteristics

The incidence of postoperative vasoplegia for our population was 28.9% (n = 55). Average age at implant was 53.1 years (range 18.6–74.3) and there were 141 (74%) males. Our cohorts were stratified into patients with and without postoperative vasoplegia. Overall demographics between groups were not significantly different, including age (p = 0.90), BMI (mean 29.9, SD 6.3, p = 0.75), race (25% African American, p = 0.88), and sex (p = 0.67) (Table 1). Further, there were no differences in estimated glomerular filtration rate (mean 67.0, SD 26.4, p = 0. 77), cardiomyopathy etiology (39.3% ischemic cardiomyopathy, p = 0.74), and indication for LVAD implant (34.8% bridge to transplant, p = 0.97).

**Table 1:** Baseline Characteristics.

|  | **No Vasoplegia** | **Vasoplegia** | **All** | **p-value** |
| --- | --- | --- | --- | --- |
|  | n = 135 | n = 55 | n = 190 |  |
| Mean Age, years (SD) | 53.2 (12.5) | 52.9 (12.3) | 53.1 (12.4) | 0.9070 |
| Mean BMI (SD) | 29.8 (6.0) | 30.1 (7.0) | 29.9 (6.3) | 0.7518 |
| African American (%) | 33 (24%) | 14 (26%) | 47 (25%) | 0.8837 |
| Male (%) | 99 (73%) | 42 (76%) | 141 (74%) | 0.6650 |
| ICM (%) | 53 (39%) | 23 (42%) | 76 (40%) | 0.7440 |
| BTT (%) | 47 (35%) | 19 (35%) | 66 (35%) | 0.9718 |
| HM3 (%) | 26 (19%) | 19 (35%) | 45 (24%) | 0.0246 |
| HM2 (%) | 57 (42%) | 12 (22%) | 69 (36%) | 0.0080 |
| HVAD (%) | 52 (39%) | 24 (44%) | 76 (40%) | 0.5137 |

### Comorbidities

Patient history of pre-operative comorbidities and conditions including hypertension, COPD, obstructive sleep apnea, diabetes mellitus, myocardial infarction, stroke, coronary artery disease, or prior deep vein thrombosis or pulmonary embolism were not significantly different between cohorts (Table 2). Further, there was no difference in patient history of prior coronary artery bypass grafting (CABG), prior percutaneous coronary intervention (PCI), and exposure to extracorporeal membrane oxygenation (ECMO). The only difference was the proportion of patients with implantable cardioverter-defibrillators (ICDs) was higher in patients without postoperative vasoplegia (82.0%) versus those with postoperative vasoplegia (66.7%, p = 0.02).

**Table 2:**
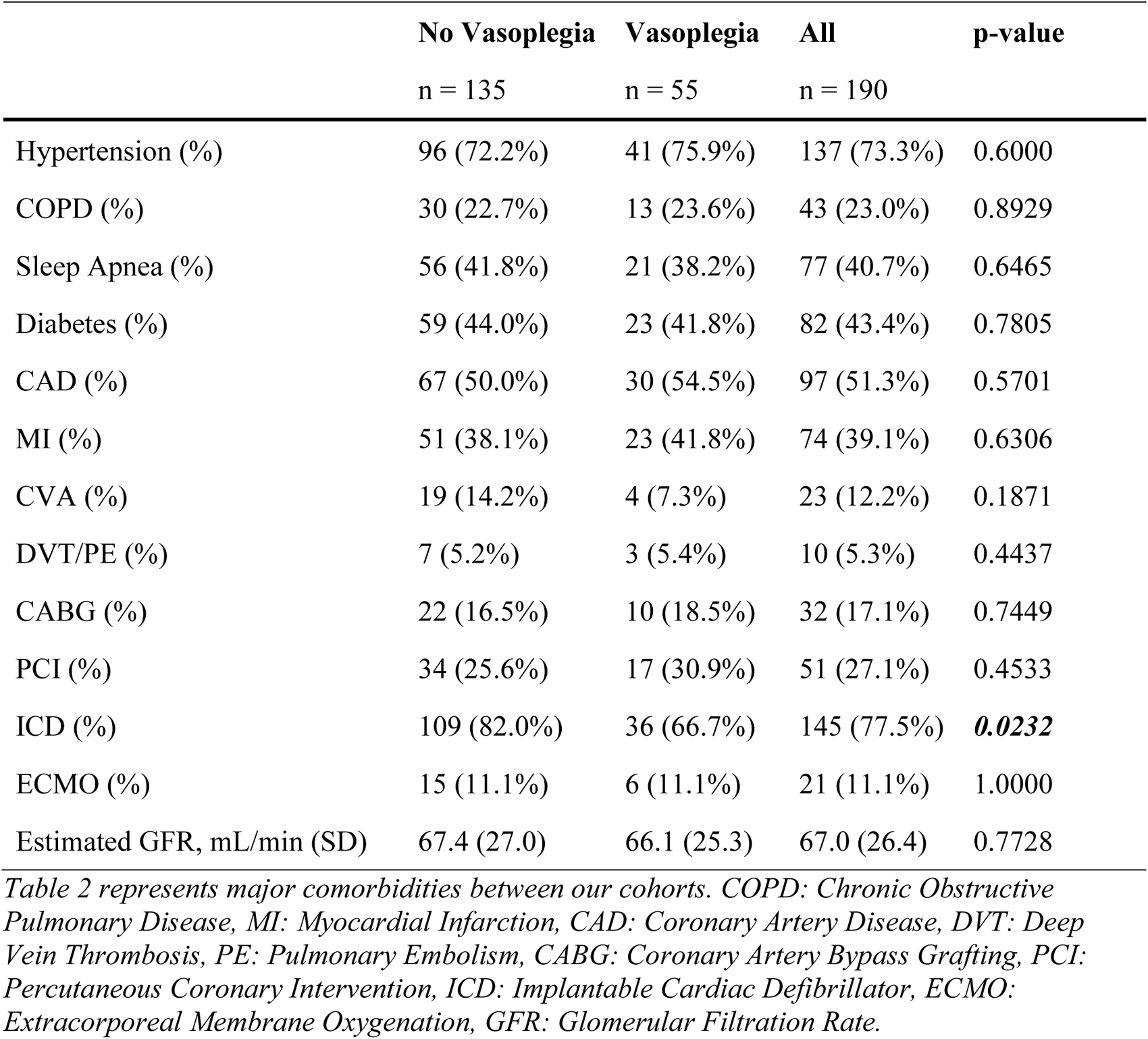
Comorbid Conditions.

### Preoperative Medications

Preoperative heart failure directed medical therapy, including renin-angiotensin axis inhibitors, combination angiotensin receptor/neprilysin inhibitors (ARNI), beta-adrenergic blockers, and diuretics were not significantly different between cohorts (Table 3). One difference in medication was patients without vasoplegia were significantly more likely to be prescribed preoperative inotropes (72.4%) compared to patients with postoperative vasoplegia (56.4%, p = 0.03).

**Table 3:**
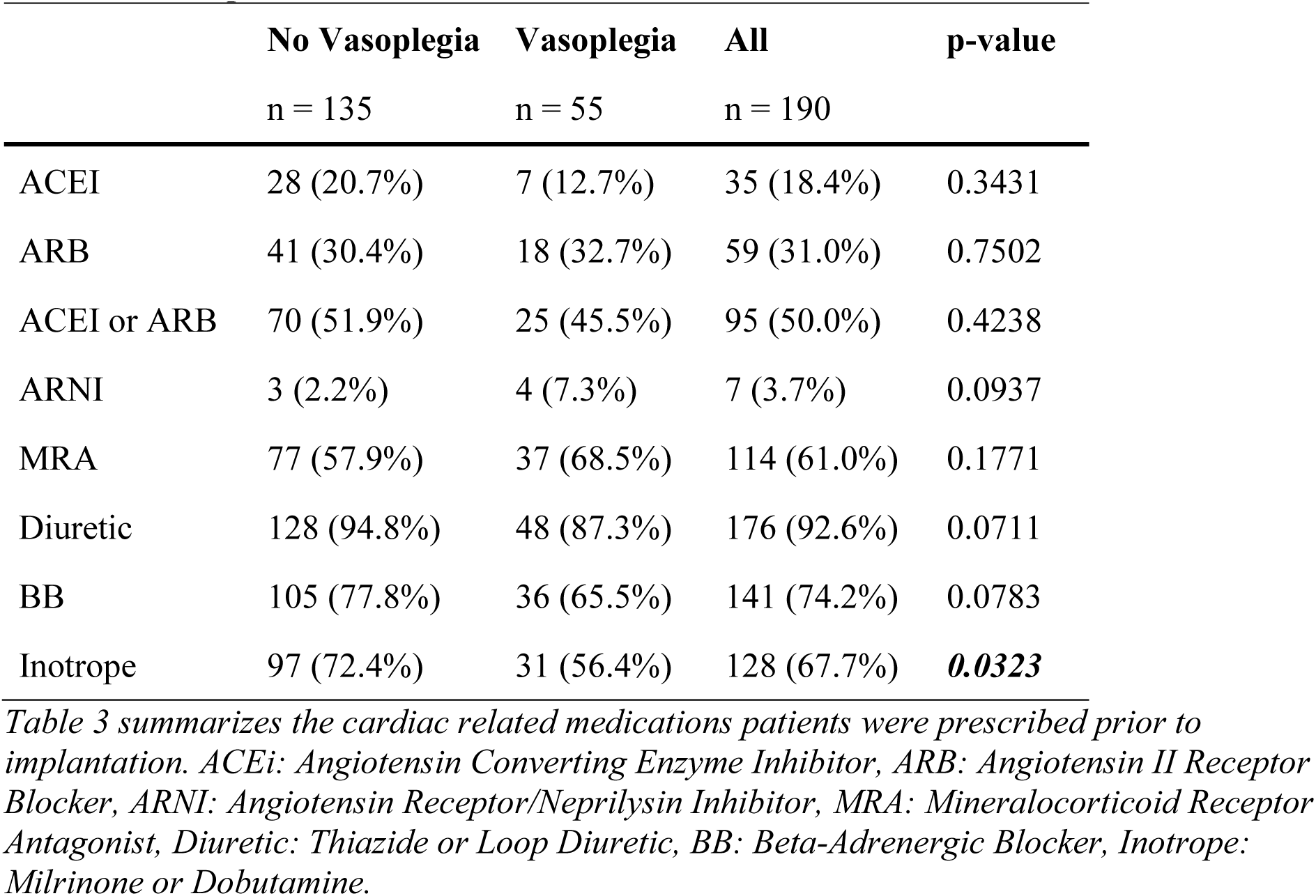
Pre-Operative Medications.

### Postoperative Blood Products

In the 48-hour period following LVAD implantation, there was no significant difference in overall blood product transfusion between cohorts (Table 4). Further, there continued to be no difference with stratification into packed red blood cells, fresh frozen plasma, and cryoprecipitate. The only difference was four patients (7.3%) with postoperative vasoplegia received postoperative platelet transfusion, while no patients received platelets in the cohort without postoperative vasoplegia (p < 0.01).

**Table 4:**
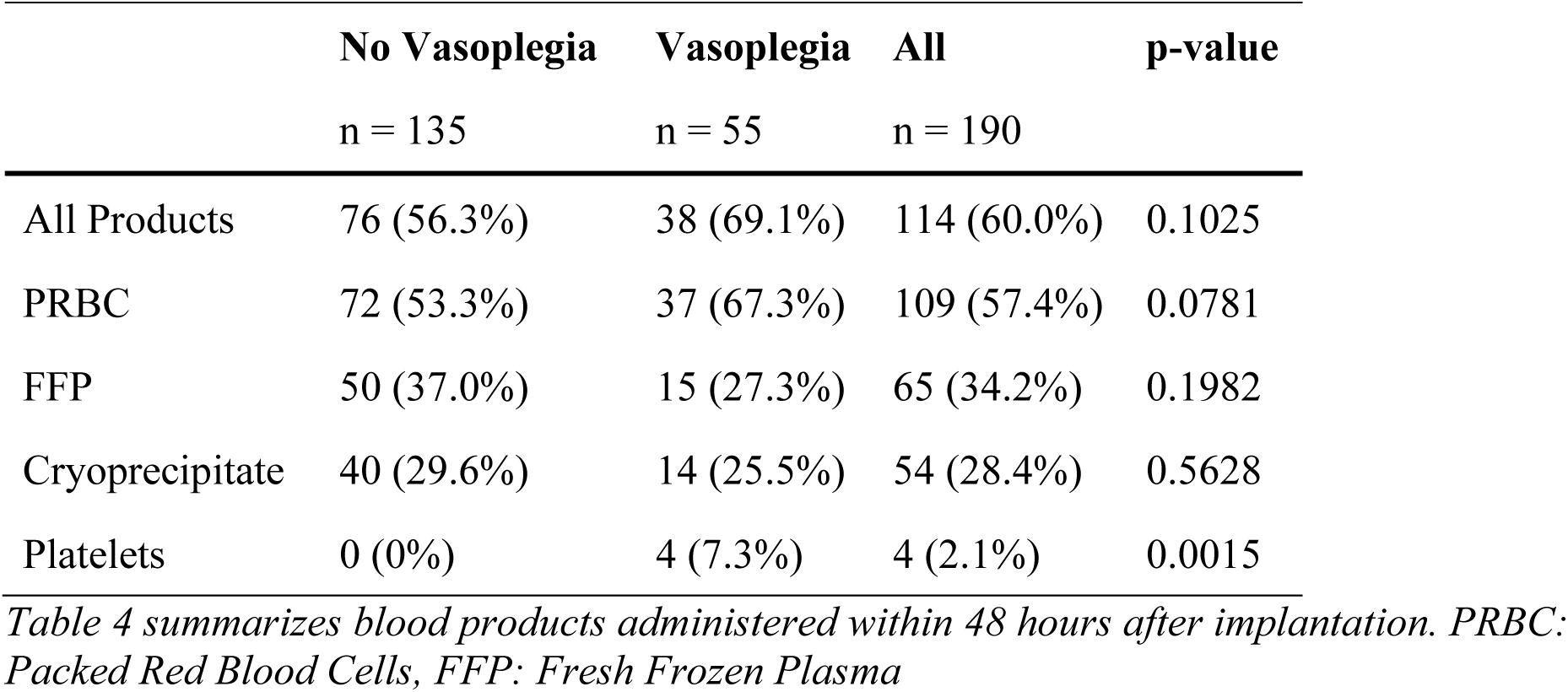
Post-Operative Blood Products.

### Outcomes

The length of intensive care unit stay in patients with vasoplegia was significantly longer (median 7 days, IQR 4–16 days) compared to patients without postoperative vasoplegia (median 5 days, IQR 3–10 days, p = 0.03). Total length of hospitalization was not significantly different between cohorts, with a median of 33.5 days for the study group (Table 5).

**Table 5:** Primary Outcomes.

|  | No Vasoplegia<br>n = 135 | Vasoplegia<br>n = 55 | All<br>n = 190 | p-value |
| --- | --- | --- | --- | --- |
| Median duration of ICU stay, days [IQR] | 5 [3, 10] | 7 [4, 16] | 6 [3, 11] | **0.0265** |
| Median duration of hospitalization, days [IQR] | 31 [20, 46] | 34 [25, 51] | 33.5 [22, 49] | 0.1651 |
| No. Early RHF (%) | 53 (39.3%) | 24 (43.6%) | 77 (40.5%) | 0.5773 |
| No. Late RHF (%) | 13 (9.6%) | 7 (12.7%) | 20 (10.5%) | 0.5281 |
| 30-day Mortality | 9 (6.7%) | 7 (12.7%) | 16 (8.4%) | 0.1725 |
| 1-year Mortality | 42 (31.1%) | 16 (29.1%) | 58 (30.5%) | 0.7839 |
| 2-year Mortality | 49 (39.2%) | 13 (32.5%) | 62 (37.6%) | 0.4463 |

Further, there was no significant difference between cohorts in developing RHF (Table 5). Overall, 40.5% (p = 0.58) of patients developed early RHF and 10.5% (p = 0.53) of patients developed late RHF (Table 5).

Finally, there was no significant difference in mortality between cohorts at 30-days, 1-year, and 2-years. Our total cohort’s 30-day mortality was 8.4 % (p = 0.17), 1-year mortality was 30.5% (p = 0.78), and 2-year mortality was 37.6% (p = 0.45).

## Discussion

Overall, we found our rate of vasoplegia at 29% to be similar to rates previously reported in literature of 5% to 45%^3–6^. Demographics and comorbid conditions were similar between cohorts, however patients with ICD implantation were less likely to experience vasoplegia. Inotropes were the only preoperative medication associated with reduced rate of vasoplegia. Interestingly, postoperative blood product administration did not have any impact on developing vasoplegia. Patients who experienced postoperative vasoplegia had a significantly longer length of ICU stay but not overall hospital length of stay. Finally, vasoplegia was not associated with developing early or late RHF or mortality.

While vasoplegia burden is of growing concern following cardiac surgery, literature thus far is equivocal about the actual mortality risk of postoperative vasoplegia. Our results contradict a similarly sized study (*n* = 252) showing higher 30-day mortality in patients with vasoplegia requiring >2 vasopressors (17.5% versus 8.4%)^8^. However, a smaller study of 24 patients showed no difference in 1-year mortality^9^.

Several cardioactive medications have been investigated as potential risk factors for vasoplegia. In CABG and orthotopic heart transplant, preoperative use of ACE-inhibitors were identified as risk factors for postoperative vasoplegia^16,26^. However, both our study and Joseph et. al. found no relationship between the use of beta-blockers or ACE-inhibitors and vasoplegia^27^.

Preoperative use of inotropes has also been associated with decreased risk for vasoplegia in patients undergoing heart transplantation in a few prior studies^12,17^. While the specific type of positive inotrope is not enumerated, certain inotropic medications, such as dopamine, have inherent vasoconstrictive properties which may explain their benefit in vasoplegic patients. However, the principal positive inotropic medication used in the present study was dobutamine, which has inherent vasodilatory properties due to its action as a beta-2 agonist. While it is primarily neutral from a vasoactive standpoint, some patients can experience transient hypotension due to vasodilation with dobutamine^28^.

While it may seem counter-intuitive, large meta-analyses have shown that positive inotropic medications, like dobutamine, especially in combination with norepinephrine have significant short-term mortality benefits in patients with septic shock. It is postulated that this combination improves microcirculatory function. Increasing evidence has shown that the microcirculatory environment may be as important in tissue perfusion regulation than macrovascular factors such as cardiac output, mean arterial pressure, and vessel tone^27^.

Further, vasoplegia extends beyond vasodilation and hypotension, causing increased vascular permeability secondary to endothelial barrier dysfunction. Mechanistically, both dobutamine and milrinone are positive inotropes via increased myocardial cyclic AMP (cAMP). Mechanistic studies have extensively shown cAMP upregulation improves vascular endothelial cell barrier function^29–32^.

However, it is also possible that given the increased cardiac output driven by positive inotropes patients can better compensate for their decreased vascular tone. Given vasoplegia is primarily defined on a basis of low blood pressure, perhaps there is a cohort of patients who have significant vasodilation consistent with vasoplegia but do not meet criteria for diagnosis as their increased cardiac output maintains their blood pressures above diagnostic thresholds. Regardless, given the short-term mortality increases in vasoplegic patients this distinction would be merely semantic.

Given these systemic and microcirculatory mechanisms, it is not unreasonable to hypothesize pre-operative inotropic medications protect endothelial cells from vasoplegic endothelial dysfunction. Thus, large, multi-institutional prospective trials into the potential benefits of positive inotropic medications for patients at risk of vasoplegia is warranted.

The principal limitation of this study is the setting of a single-institutional retrospective analysis and thus results may not be generalizable to other centers. However, this is true of most current literature regarding vasoplegia post-operatively. As alluded to above, vasoplegia lacks a consensus clinical definition, and a wide spectrum of criteria have been used in previous literature. While our definition is similar, the wide variety of definitions in the literature make true comparison of outcomes difficult. Additionally, inherent to retrospective studies, variables were not recorded with the intent for research and thus some variables were unavailable or incomplete for analysis. We also contend that there may be potential variation in the treatment of hypotension amongst providers, however given the consistency of treatment modality for vasoplegic patients we contend that this variation would provide little effect, and this is a limitation that would be consistent across the literature.

Further, ICDs were more prevalent in our cohort of patients without postoperative vasoplegia. This finding potentially highlights an underlying selection bias where patients with ICDs and inotropes may have received more pre-operative heart failure specific care and thus were more optimized ahead of surgery. Finally, given the wide range of objective vasoplegia definitions, we opted to include physician discretionary criterion as a substitute to include all patients who were treated for vasoplegia by our heart failure specialists.

## Conclusion

Vasoplegia is a significant risk following cardiac surgery, particularly implantation of left ventricular assist devices. While short term mortality is high, mortality is equivocal suggesting that better risk assessment and treatment modalities are needed. One such strategy could involve the assessment of pre-operative positive inotropic medications and risk of developing vasoplegia post-operatively, which showed promise in the present retrospective analysis. Further work is needed to investigate the potential treatment efficacy of these drugs prospectively.

## Data Availability

Data available in the manuscript is available for review upon request

## Non-standard Abbreviations and Acronyms

|  |  |
| --- | --- |
| **ARNI** | Angiotensin Receptor/Neprilysin Inhibitor |
| **BB** | Beta-Adrenergic Blocker |
| **BTT** | Bridge to Transplant |
| **CVP** | Central Venous Pressure |
| **DT** | Destination Therapy |
| **HM2** | HeartMate 2 |
| **HM3** | HeartMate 3 |
| **HVAD** | HeartWare Ventricular Assist Device |
| **ICM** | Ischemic Cardiomyopathy |
| **MRA** | Mineralocorticoid Receptor Antagonist |
| **OHT** | Orthotopic Heart Transplant |
| **PRBC** | Packed Red Blood Cells |
| **RHF** | Right Heart Failure |

